# Genomic insights into the mechanism of NK3R antagonists for treatment of menopausal vasomotor symptoms

**DOI:** 10.1101/2022.05.25.22275501

**Authors:** Katherine S. Ruth, Robin N. Beaumont, Jonathan M. Locke, Jessica Tyrrell, Carolyn J. Crandall, Gareth Hawkes, Timothy M. Frayling, Julia K. Prague, Kashyap A. Patel, Andrew R. Wood, Michael N. Weedon, Anna Murray

## Abstract

**Background:** Menopausal vasomotor symptoms (VMS) significantly impact women’s quality of life, and whilst hormone replacement therapy (HRT) is effective, it is not appropriate for all. We aimed to identify new drug targets for VMS and understand reasons for HRT use through genomic analyses.

**Methods:** In up to 153,152 women from UK Biobank, a population-based cohort, we performed a genome-wide association study (GWAS) of VMS derived from linked primary-care records and cross-sectional self-reported data. In a subset of this cohort (n=39,356), we analysed exome-sequencing data to test the association of rare deleterious genetic variants with VMS. Finally, we used Mendelian randomisation analysis to investigate the reasons for HRT use and whether these changed over time.

**Findings:** Our GWAS identified a genetic signal near the gene encoding NK3R (*TACR3*) associated with a lower risk of VMS (OR=0·85 (95% CI 0·82,0·87) per AT allele, P=1·1×10^-26^), which was consistent with previous studies. However, rare genetic variants predicted to reduce functional NK3R levels were not associated with VMS (P=0·9), though did delay puberty (P=9×10^-11^). Younger menopause age was causally-associated with greater HRT use before 2002 but not after.

**Interpretation:** Using genomics we demonstrate that changed HRT use since the early 2000s reflects a switch from preventing post-menopausal complications to primarily treating VMS. We provide support for *TACR3* in the genetic basis of VMS but unexpectedly find that rare genomic variants predicted to lower NK3R levels did not modify VMS, despite the proven efficacy of NK3R antagonists, suggesting that further biological understanding could benefit therapeutic efficacy.

**Funding:** Cancer Research UK and UKRI.

**Research in context:** *Evidence before the study:* *In vivo* studies of animal models and clinical studies in humans have determined that menopausal vasomotor symptoms (VMS) result from increased neurokinin B (NKB) signalling via the neurokinin 3 receptor (NK3R) in response to decreased circulating oestradiol levels. Recent Phase II clinical trials have demonstrated the efficacy of NK3R antagonists in reducing VMS. A previous GWAS in 17,695 women identified a genetic signal at the *TACR3* locus (which codes for NK3R) as associated with VMS. The locus was also genome-wide significant in a GWAS of oestrogen-replacement use (15,305 cases) derived from self-reported medications in UK Biobank.

*Added value of this study:* This study represents a novel approach to analysing the rarely captured phenotype of VMS, since few population-based cohorts have asked about menopausal symptoms. To the best of our knowledge, this is the first analyses of VMS identified from linked primary care health records. Literature searches of published papers and codelists have not identified any previous studies of VMS in primary care data. The replication of the known GWAS signal for VMS provides a validation of the coding of this phenotype from primary care data. This is the largest genomic study of VMS currently carried out (92,028 women). Our current analyses are limited by the availability of primary care linked data in ∼45% of the UK Biobank cohort and are based on exome sequencing in 200,000 women. Recently released exome data for the full cohort and further releases of primary care linked data in UK Biobank will allow us to re-visit these analyses further.

*Implications of all the available evidence:* Our analyses of rare coding variation in *TACR3* identified an intriguing difference that requires further study; while NK3R antagonist drug treatment reduces VMS, women carrying rare genetic variants resulting in reduced NK3R levels were no less likely to experience VMS. Our genome-wide analyses replicate the genetic signals for VMS at the *TACR3* gene locus, however we were unable to unequivocally identify *TACR3* as the causal gene at this locus. We suggest that the effect of the common genetic variant on reducing VMS may be through as yet uncharacterised regulatory pathways, and that complete inhibition of NK3R signalling is required to eliminate (rather than reduce) VMS.

## Introduction

Vasomotor symptoms (VMS) in postmenopausal women include hot flashes and night sweats, which can have debilitating and long-lasting effects on quality of life, with 10% of women experiencing VMS for up to 12 years.^1^ Hormone replacement therapy (HRT) is the most effective treatment for VMS around menopause but its popularity amongst the public and health professionals declined following the publication of adverse outcome data^2^ and it is contraindicated in some women, such as those who have had breast cancer. Consequently, there is great interest in identifying non-oestrogen drug treatments for VMS.

VMS result from dysfunction of thermoregulation by the hypothalamus and autonomic thermoregulatory system caused by increased neurokinin B (NKB) /neurokinin 3 receptor (NK3R; also known as the neuromedin-K receptor or tachykinin receptor 3) signalling in response to decreased circulating oestradiol levels, as shown *in vivo* in animal models.^3,4^ Four Phase 2 clinical trials have demonstrated that administration of a NK3R receptor antagonist results in a clinically significant reduction in hot flushes comparable to that achieved by HRT.^5–8^ Such NK3R receptor antagonists are currently in Phase 3 clinical trials and have not yet come to market.^9^ Neurokinin signalling also plays an important role in controlling the onset of puberty^10^, with genetic variants that knock out the *TACR3* gene (which encodes NK3R) resulting in pubertal failure in homozygous carriers^11,12^ and delayed puberty in heterozygous carriers.^13^ Additionally, population-based genome-wide association studies (GWAS) have identified several more common genetic variants in or near the *TACR3* gene associated with normal variation in age at menarche.^14^

We aimed to increase understanding of the mechanism of VMS and to identify other novel treatment targets using a combination of GWAS and exome sequencing. A previous GWAS in 17,695 women identified a signal in *TACR3*.^15^ Our study in UK Biobank (N=153,152) included nearly 10 times more women than this initial GWAS and we have used a combination of linked primary care health records and self-reported questionnaire data to capture VMS by different methods to validate our findings.^16,17^

## Methods

### Phenotype definitions

Between 2006-2010, UK Biobank recruited over 500,000 individuals aged 37-73 years from across the UK who answered detailed questions about themselves, had measurements taken and provided blood, urine and saliva sample samples, and for whom linked health records data are available.^17^ We identified women with VMS for inclusion in our primary analyses from linked primary care records, which are available for ∼45% of the cohort and capture participants’ contact with health care professionals working at UK general practices (family physicians) over their lifetime to ∼2017. Cases (n=14,261) were women with one or more clinical events in the linked primary care records (e.g. symptoms, history or diagnosis) containing any of 33 relevant codes for menopausal VMS (Supplementary Methods; Supplementary Table 1). Controls (n=77,767) were women who were included in the linked primary care records (identified through having a primary care registration record without a relevant VMS code), and aged 50 and over at the baseline UK Biobank assessment and so were likely to be experiencing the menopause transition and thus at risk of VMS.

For secondary analyses we derived proxy phenotypes for VMS based on self-reported HRT use (Supplementary Methods, Supplementary Table 2). To explore the effect of the publication of adverse health outcomes relating to HRT, we further subdivided the ever taking HRT group into those who took HRT before and after 2002 to reflect the year of publication of the initial findings of the Women’s Health Initiative HRT trial^18^ and estimated the proportion of women taking HRT before and after 2002 (Supplementary Methods).

### Genome-wide association study analyses

Genotyping of the UK Biobank study was performed centrally^16^. Two genotyping arrays with over 95% common marker content were used to genotype the individuals; the Affymetrix Axiom UK Biobank array (∼450,000 individuals) and the UKBiLEVE array (∼50,000 individuals). This dataset underwent extensive central quality control and genotype imputation using 1000 Genomes Phase 3/UK10K and Haplotype Reference Consortium reference panels. We based our study on 451,099 individuals who we identified as being of European descent, as described previously.^19^

We carried out genome-wide analyses using BOLT-LMM v2.3^20^, which uses a mixed linear model to account for relatedness and ethnicity. We tested 17 million genetic variants with minor allele frequency (MAF) >0·1% and imputation quality >0·3. Association testing was based on an additive model and was adjusted for genotyping chip and release of the data, recruitment centre and age (Supplementary Methods). Quantitative traits were inverse-rank normalised, to ensure that residuals were normally distributed. Genome-wide significant variants had *P*<5×10^-8^. For case-control traits we transformed effect sizes (*β*) on the quantitative scale to odds ratios (using the transformation ln(OR)=*β*/(*μ*×(1-*μ*)), where *μ* is the fraction of cases) and confirmed associations using a Fisher’s exact test (Supplementary Methods). We performed case-control matched analyses of HRT use to account for different age distributions (Supplementary Methods). Independent signals were more than 500kb from the next most significant variant for the same phenotype and in linkage disequilibrium (LD) *r*^*2*^<0.5 with signals for other VMS phenotypes. Additionally, to identify further independent signals at the *TACR3* locus, we carried out conditional analysis by re-running the VMS GWAS including the genotype at the lead variant as a covariate.

We meta-analysed the results of the VMS GWAS with summary statistics from the European cohorts (GARNET and WHIMS) of Crandall et al 2017 (2,747 cases and 5,440 controls).^15^ Inverse-variance weighted meta-analyses were carried out in METAL^21^ with genomic-control correction applied and we included variants with MAF >0·1% and imputation quality >0·3.

### Annotation of GWAS signals

We tested whether our GWAS signals were associated with age at menarche and age at menopause in UK Biobank by conducting GWAS (method as described) using phenotypes consistent with those used in ReproGen consortium analyses (n=106,048 for age at menopause; n=236,807 for age at menarche).^14,22^ We also calculated LD between our GWAS signals and previously published signals for menarche and menopause timing.^14,22^ LD was calculated for the regions around the HRT signals from best guess genotypes for 1000 Genomes Phase 3/HRC imputed variants in ∼340,000 unrelated UK Biobank participants of white British ancestry using PLINK v1.9.^23^ To identify variants with functional or regulatory consequences, we looked up variants in LD with our lead genetic variant in Variant Effect Predictor (build 38)^24^, Ensembl (Human (GRCh38.p13 release 100)^25^, HaploReg v4.1^26^ and goDMC (http://mqtldb.godmc.org.uk/). We investigated eQTLs in r^2^>0·8 with the top hits in PsychENCODE^27^ and GTEx v8 (https://www.gtexportal.org/home). We calculated the distance of variants in r^2^>0·8 with the lead genetic variant to canonical and alternative splice sites for *TACR3* using Intropolis.^28^ Manhattan and quantile-quantile plots were produced in R using the package “qqman”.^29^ LocusZoom *v1*.4^30^ was used to plot the association statistics at individual loci.

### Exome-wide analyses

We carried out analyses of association of VMS and age at menarche with rare variants using exome sequencing data available in ∼200,000 people from UK Biobank (Supplementary Methods).^31^ Variants in CCDS transcripts were annotated using Variant Effect Predictor^24^ and we identified loss-of-function (LOF) variants (stop-gain, frameshift, or abolishing a canonical splice site (-2 or +2 bp from exon, excluding the ones in the last exon)) deemed to be high confidence by LOFTEE (https://github.com/konradjk/loftee). We conducted gene-burden analyses using SAIGE-GENE^32^ for age at menarche and REGENIE^33^ for VMS, which account for relatedness and ethnicity and are suitable for analyses with unbalanced case:control ratios. We included recruitment centre and age at baseline as covariates and tested the association of variants with MAF<0.001 in each gene in aggregate as well as individually. For each gene, we present results for the transcript with the smallest burden p-value.

### Tests of associations of variants in the TACR3 *region*

We investigated whether puberty timing genetic variants in/near the *TACR3* gene were also associated with VMS. These variants included a rare protein truncating variant which in homozygous state leads to hypogonadotropic hypogonadism and in heterozygous state to delayed menarche (rs144292455 C>T p.W275X, chr4:104577415, MAF=0·06%)^13,34^ and five genetic variants associated with age at menarche in GWASs (rs55784701 (chr4:104247262), rs3733632 (chr4:104640935), rs62342064 (chr4:104665972), rs115260227 (chr4:104774698), rs17035311 (chr4:106066293), smallest MAF 1·3%) within ∼1·5Mb of *TACR3*.^14^ Genotypes were extracted from imputed data except for rs144292455, which is a rare variant and thus poorly imputed; for this variant we used directly genotyped data, which have previously been shown to be reliable.^35^ We tested the associations of these variants individually, in combination and when adjusted for genotype of our lead VMS genetic variant using regression analyses. We performed logistic (binary traits) and linear (quantitative traits) regression in Stata v14.0/v16.0 in 379,768 unrelated individuals of European descent.^36^ We regressed outcomes on genotype including the covariates genotyping chip and release of genotype data, recruitment centre, age and the first five genetic principal components (generated as described previously^36^).

### Mendelian randomisation analyses

Given the overlap in the genetic variants identified for HRT use and age at natural menopause, we used Mendelian randomisation (MR) analyses to test whether earlier age at menopause was the cause of women taking HRT, or just correlated with HRT use. We constructed a genetic instrument for the exposure age at menopause from 56 published genetic variants discovered in a meta-analyses of ∼70,000 women that was independent of UK Biobank data.^22^ In each MR test we assessed a number of widely used methods including inverse-variance weighted MR (IVW MR) and those more robust to pleiotropy (weighted median MR, penalised weighted median MR and MR–Egger^37,38^) to address the assumption that alleles that influence the exposure do not influence the outcome via any pathway other than through the exposure. We used IVW MR as our primary analysis method and interpreted directionally-consistent results across the various methods as strengthening our causal inference.

### Heterogeneity in effects of genetic variants before and after 2002

Following the publication of data on adverse effects of HRT in 2002, we hypothesized that the primary reasons why women took HRT changed before and after this date, and that these differences may not have been solely due to a change in preference of women and health professionals and would be reflected in differences in genetic associations by time. To test this, we compared the direction of effect of the lead variant for the 15 signals on ever using HRT before and after 2002. We compared effect estimates from the GWAS of HRT use before and after 2002 by calculating a heterogeneity chi-squared *P*-value using the package “metan” in Stata v14.0/v16.0. We carried out Mendelian randomisation (as described) to test the association of genetically-instrumented age at menopause with HRT use before and after 2002.

## Results

We identified 14,261 women with VMS and 77,767 controls from linked primary care records in UK Biobank for inclusion in our genome-wide association study (GWAS) of VMS (Table 1). From our GWAS we identified a single independent genetic signal (lead variant rs34867104) associated with lower odds of having VMS ((OR=0·78 (95% CI 0·74,0·82) per AT allele; allele frequency (AF) = 5·5%; *P*=1·7×10^-20^) (Supplementary Figure 1, Figure 1). Meta-analysis of the UK Biobank VMS GWAS with that from Crandall et al 2017^15^ replicated the same single genetic signal in the *TACR3* gene (OR=0·76 per A allele (95% CI 0·72, 0·80; P=3·7×10^-27^) (Table 2). No further independent signals at *P*<5×10^-8^ were identified when we performed GWAS conditioning on the genotype at the lead variant (rs34867104). *In silico* analyses found little evidence to suggest a biological mechanism for the GWAS signal. There were no non-synonymous variants (with effects on protein sequence) strongly correlated with the signal (LD r^2^>0·8), and there was little evidence for an effect on gene expression, regulation or splicing (Supplementary Results). However, based on clinical studies that have demonstrated that NK3R antagonists reduce VMS^5,39,40^, *TACR3* is the most likely causal gene in this region.

**Table 1.** Descriptive statistics for demographic and reproductive characteristics of women included as cases and controls in GWAS of VMS phenotypes.

|  | Vasomotor symptoms |  |  |  | Ever taken HRT |  |  |  |
| --- | --- | --- | --- | --- | --- | --- | --- | --- |
|  | Cases |  | Controls |  | Cases |  | Controls |  |
|  | N | Mean (SD) / % (n) | N | Mean (SD) / % (n) | N | Mean (SD) / % (n) | N | Mean (SD) / % (n) |
| Age | 14,261 | 56.08 (7.37) | 77,767 | 60.55 (5.39) | 72,622 | 61.82 (5.09) | 80,686 | 59.6 (5.81) |
| Body mass index | 14,209 | 27.25 (5.03) | 77,489 | 27.31 (5.11) | 72,371 | 27.04 (4.81) | 80,417 | 27.00 (5.16) |
| Age at menarche | 13,832 | 12.95 (1.6) | 75,335 | 12.93 (1.57) | 70,614 | 12.94 (1.57) | 78,102 | 12.94 (1.54) |
| Age at natural menopause | 4,956 | 50.17 (4) | 42,920 | 50.47 (3.91) | 33,250 | 49.53 (4.33) | 72,629 | 50.7 (3.74) |
| Number of births | 14,253 | 1.80 (1.15) | 77,719 | 1.90 (1.16) | 72,583 | 1.91 (1.14) | 80,640 | 1.85 (1.18) |
| Number of pregnancies | 13,977 | 2.31 (1.56) | 76,437 | 2.34 (1.54) | 71,437 | 2.39 (1.54) | 79,382 | 2.29 (1.55) |
| Townsend deprivation index | 14,241 | -1.51 (2.9) | 77,673 | -1.57 (2.89) | 72,544 | -1.57 (2.91) | 80,607 | -1.62 (2.87) |
| Smoking status | 14,087 |  | 76,901 |  | 71,709 |  | 79,915 |  |
| - never |  | 56.2% (7,912) |  | 58.6% (45,057) |  | 53.1% (38,102) |  | 61.8% (49,351) |
| - previous |  | 33.8% (4,768) |  | 34.1% (26,208) |  | 38.8% (27,845) |  | 31.3% (25,023) |
| - current |  | 10.0% (1,407) |  | 7.3% (5,636) |  | 8.0% (5,762) |  | 6.9% (5,541) |
| Vasomotor symptoms | 14,261 |  | 77,767 |  | 33,015 | 16.8% (5,533) | 36,106 | 7.5% (2,717) |
| Ever taken HRT | 8,250 | 67.1% (5,533) | 60,871 | 45.1% (27,482) | 72,622 |  | 80,686 |  |
| Used HRT before 2002 | 6,779 | 59.9% (4,062) | 55,748 | 40.1% (22,359) | 57,740 |  | 80,686 |  |
| Used HRT after 2002 | 3,625 | 25.0% (908) | 35,304 | 5.4% (1,915) | 6,640 |  | 80,686 |  |

*Notes: All variables are at baseline except for age at menopause, which was calculated from the the most recent available visit. Ever taken HRT was defined in postmenopausal women and was identified by the answer to: "Have you ever used hormone replacement therapy (HRT)?" For categorical variables, N is the total number of cases or controls with information; Age at menarche excludes values <9 and >17 years; Age at natural menopause excludes values <40 and >60 years. Townsend deprivation scores range from negative values to positive values with smaller scores indicating less material deprivation.*

**Table 2.**
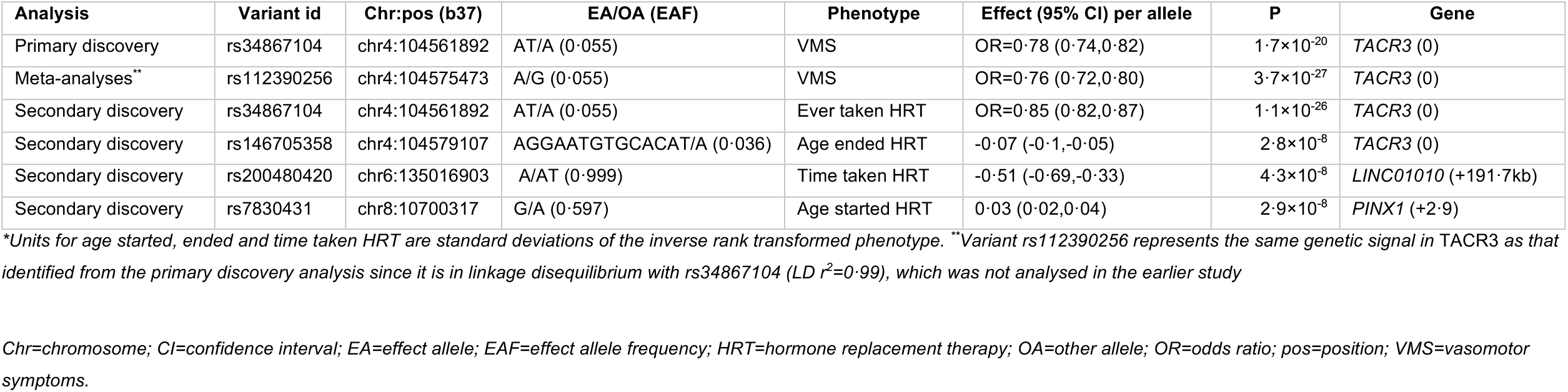
Genetic signals for VMS from primary GWAS analysis and secondary analyses of HRT proxy phenotypes.

| Analysis | Variant id | Chr:pos (b37) | EA/OA (EAF) | Phenotype | Effect (95% CI) per allele | P | Gene |
| --- | --- | --- | --- | --- | --- | --- | --- |
| Primary discovery | rs34867104 | chr4:104561892 | AT/A (0.055) | VMS | OR=0.78 (0.74,0.82) | $1.7 \times 10^{-20}$ | TACR3 (0) |
| Meta-analyses** | rs112390256 | chr4:104575473 | A/G (0.055) | VMS | OR=0.76 (0.72,0.80) | $3.7 \times 10^{-27}$ | TACR3 (0) |
| Secondary discovery | rs34867104 | chr4:104561892 | AT/A (0.055) | Ever taken HRT | OR=0.85 (0.82,0.87) | $1.1 \times 10^{-26}$ | TACR3 (0) |
| Secondary discovery | rs146705358 | chr4:104579107 | AGGAATGTGCACAT/A (0.036) | Age ended HRT | -0.07 (-0.1,-0.05) | $2.8 \times 10^{-8}$ | TACR3 (0) |
| Secondary discovery | rs200480420 | chr6:135016903 | A/AT (0.999) | Time taken HRT | -0.51 (-0.69,-0.33) | $4.3 \times 10^{-8}$ | LINC01010 (+191.7kb) |
| Secondary discovery | rs7830431 | chr8:10700317 | G/A (0.597) | Age started HRT | 0.03 (0.02,0.04) | $2.9 \times 10^{-8}$ | PINX1 (+2.9) |

\*Units for age started, ended and time taken HRT are standard deviations of the inverse rank transformed phenotype. \*\*Variant rs112390256 represents the same genetic signal in TACR3 as that identified from the primary discovery analysis since it is in linkage disequilibrium with rs34867104 ( $LD\ r^2=0.99$ ), which was not analysed in the earlier study
Chr=chromosome; CI=confidence interval; EA=effect allele; EAF=effect allele frequency; HRT=hormone replacement therapy; OA=other allele; OR=odds ratio; pos=position; VMS=vasomotor symptoms.

**Figure 1.**
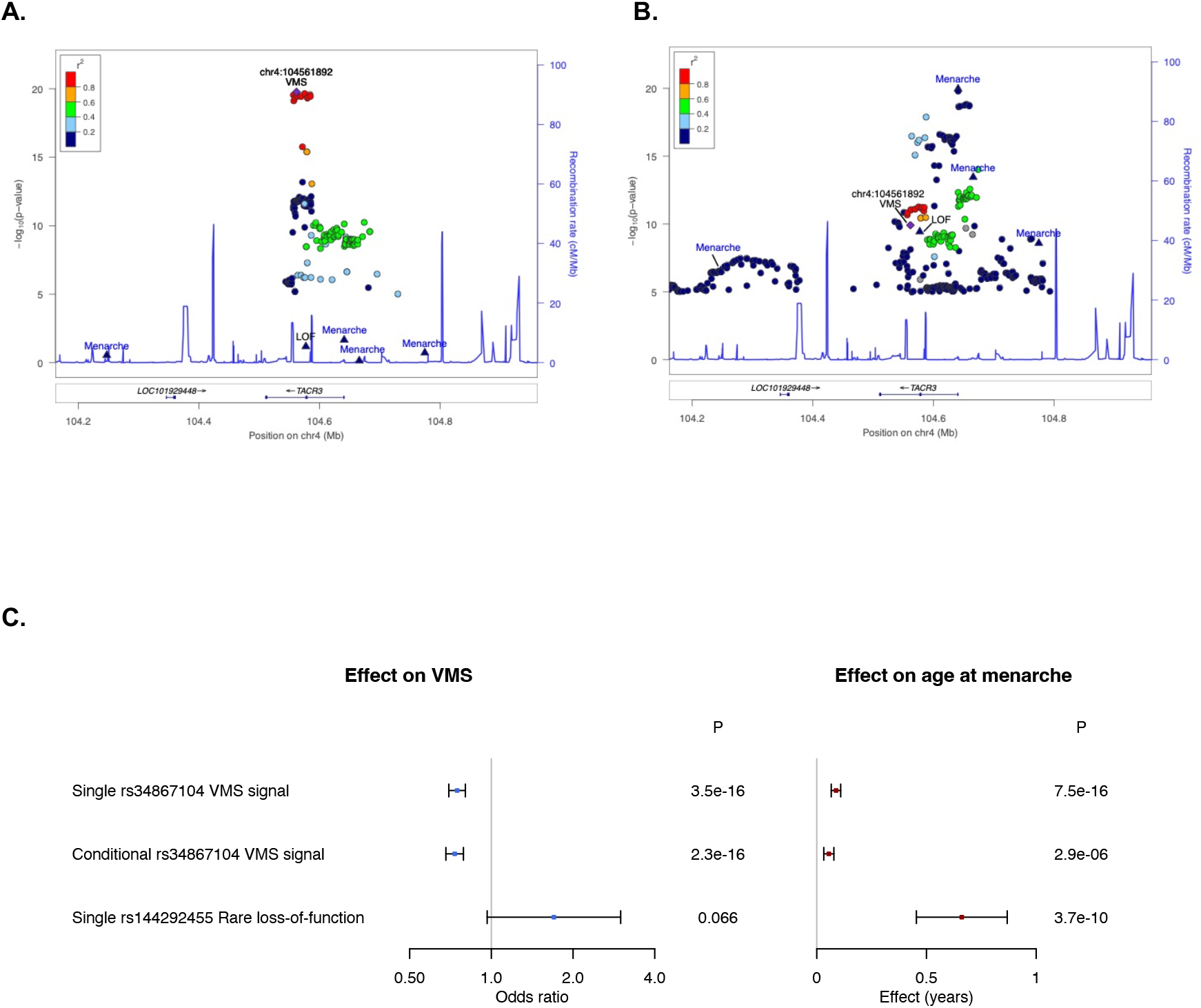
Genetic associations at the *TACR3* region: (A) Associations with VMS in UK Biobank. GWAS signals for age at menarche and the *TACR3* LOF allele (rs144292455 C>T) were not associated with VMS at P<5×10^-8^ and show little correlation with the VMS signal (LD r^2^<0.2); **(B) Associations with age at menarche in published GWAS**. GWAS signals for age at menarche and the *TACR3* LOF allele (rs144292455 C>T) were strongly associated with age at menarche. The VMS lead variant shows association at P<5×10^-8^ with age at menarche in this univariate analysis though was not identified as an independent signal in published age at menarche GWAS. **(C) Univariate and conditional association analyses in UK Biobank**. The VMS lead variant was associated with age at menarche at P<5×10^-8^ in univariate analyses (“Single”) but this association attenuated in analysis adjusting for the genotypes of the age at menarche GWAS signals (“Conditional”). The *TACR3* LOF allele (rs144292455 C>T) was strongly associated with later age at menarche but not VMS. *Notes: Variants shown are within +/-400kb of rs34867104, the lead variant for VMS, and LD r^2^ shown is with rs34867104. Labelled variants are four variants associated with age at menarche within this region (triangles)^14^, VMS (purple diamond) and also the LOF variant rs144292455 (triangle). For clarity, other variants with P=1×10^-5^ are not shown. Association statistics for rs144292455 were calculated in directly genotyped data in UK Biobank whereas all other statistics are from GWAS of imputed data. Association statistics for age at menarche GWAS are from published ReproGen meta-analyses excluding 23andMe^14^.*

Using exome sequencing data available for 6,280 women with VMS and 33,076 controls we investigated the effect of reduced levels of TACR3 protein on VMS using genetic variants in *TACR3* predicted with high confidence to result in no protein product (i.e. LOF variants). We tested the association with VMS of a *TACR3* LOF allele rs144292455 C>T p.W275X (MAF=0.06%) previously reported as associated with delayed menarche^13,34^ and all rare (MAF<0·1%) LOF variants in *TACR3* in aggregate. There was little evidence of an association between *TACR3* LOF and VMS (rs144292455 *P*=0·6, 8/41 carriers were VMS cases vs 6,272/39,315 non-carriers; aggregate LOF burden *P*=0·9, 8/49 carriers were VMS cases vs 6,272/39,307 non-carriers), though *TACR3* LOF was associated with delayed age at menarche (rs144292455 *P*=5·0×10^-9^, 105 carriers vs 98,386 non-carriers; aggregate LOF burden *P*=8·6×10^-11^, 128 carriers vs 98,363 non-carriers) (Supplementary Table 3). Associations of rs144292455 with VMS and age at menarche were consistent when we repeated the analysis in a larger sample of directly genotyped chip data (18/79 carriers were VMS cases vs 11,832/76,335 non-carriers) (Supplementary Table 4). We further confirmed the independence of the VMS GWAS signal (rs34867104) and rs144292455 through conditional analyses (Supplementary Table 4). Several GWAS signals for age at menarche are in the *TACR3* region, however these have a different underlying genetic basis to the signal for VMS (Figure 1, Supplementary Results and Supplementary Table 5). Therefore, our genetic analyses suggest that individuals with loss of *TACR3* have later puberty but do not have fewer VMS.

Next we increased our sample size by employing HRT use as a proxy phenotype for VMS, identifying variant rs34867104 as the most strongly associated signal for HRT use (Table 2) (OR=0·85 (95% CI 0·82,0·87) per AT allele, P=1·1×10^-26^), with consistent results in age-matched sensitivity analyses (OR=0·83 (95% CI 0·80,0·86)). Our secondary analyses resulted in a further 14 independent genetic signals associated with HRT phenotypes that did not reach genome-wide significance in the VMS GWAS (all P>0·05) (Supplementary Results, Supplementary Table 6). However, 11 of the 14 signals represented associations with HRT use as a consequence of early menopause; 10 signals were associated with menopause timing at P<5×10^-8^ in the largest genome-wide meta-analysis of age at natural menopause ^41^, with a further signal associated (P<5×10^-8^) in UK Biobank (Supplementary Table 6). Furthermore, Mendelian randomisation analyses showed that menopause timing is causally associated with HRT use, with a one-year genetically-instrumented earlier age at menopause strongly associated with higher odds of taking HRT (OR=1·10 (1·08,1·12), P=1×10^-18^) (Supplementary Table 7). In contrast, the VMS signal at *TACR3* (rs34867104) was not associated with menopause timing (Supplementary Table 6) and the genetic instrument for menopause timing showed little association with VMS (IVW MR P>0·05). Three additional genetic signals near *C3orf43, LINC01010* and *PINX1* were not associated with menopause timing and represent putative VMS signals highlighted by our proxy GWAS (Supplementary Table 6).

Following the publication of adverse effects of HRT in 2002, we hypothesized that only women with severe VMS took HRT, considering the benefit to outweigh the risk, and that these differences will be reflected in genetic associations. In UK Biobank we identified a change in the proportion of women using HRT from 49% before 2002 to 14% after 2002. By testing the association of genetically-instrumented age at menopause with HRT use before and after 2002, we found that a one-year genetically-predicted earlier menopause raised the odds of HRT use before 2002 (OR=1·12, 95% CI=1·10,1·13), but not after 2002 (OR=0·98, 95% CI=0·95,1·00) (Figure 2), with concordant results in age-matched sensitivity analyses (Supplementary Table 7). Consistent with this finding, the signals that were associated with age starting HRT as a result of menopause timing had opposite effects on HRT use before and after 2002 (Supplementary Table 8). In contrast, the genetic signal in *TACR3* (rs34867104) was associated with lower odds of using HRT both before and after 2002 (before 2002, OR=0·85 per AT allele (95% CI 0·83,0·88), *P*=1·6×10^-21^; after 2002, OR=0·83 per AT allele (95% CI 0·77,0·89), *P*=1·4×10^-6^; P test of heterogeneity=0·48). Two of the putative VMS signals (rs201598433 near *C3orf43* and rs200480420 near *LINC01010*) also showed little evidence of heterogeneity in effects pre/post 2002 (Supplementary Table 8), strengthening the evidence that they are candidate VMS signals.

**Figure 2.**
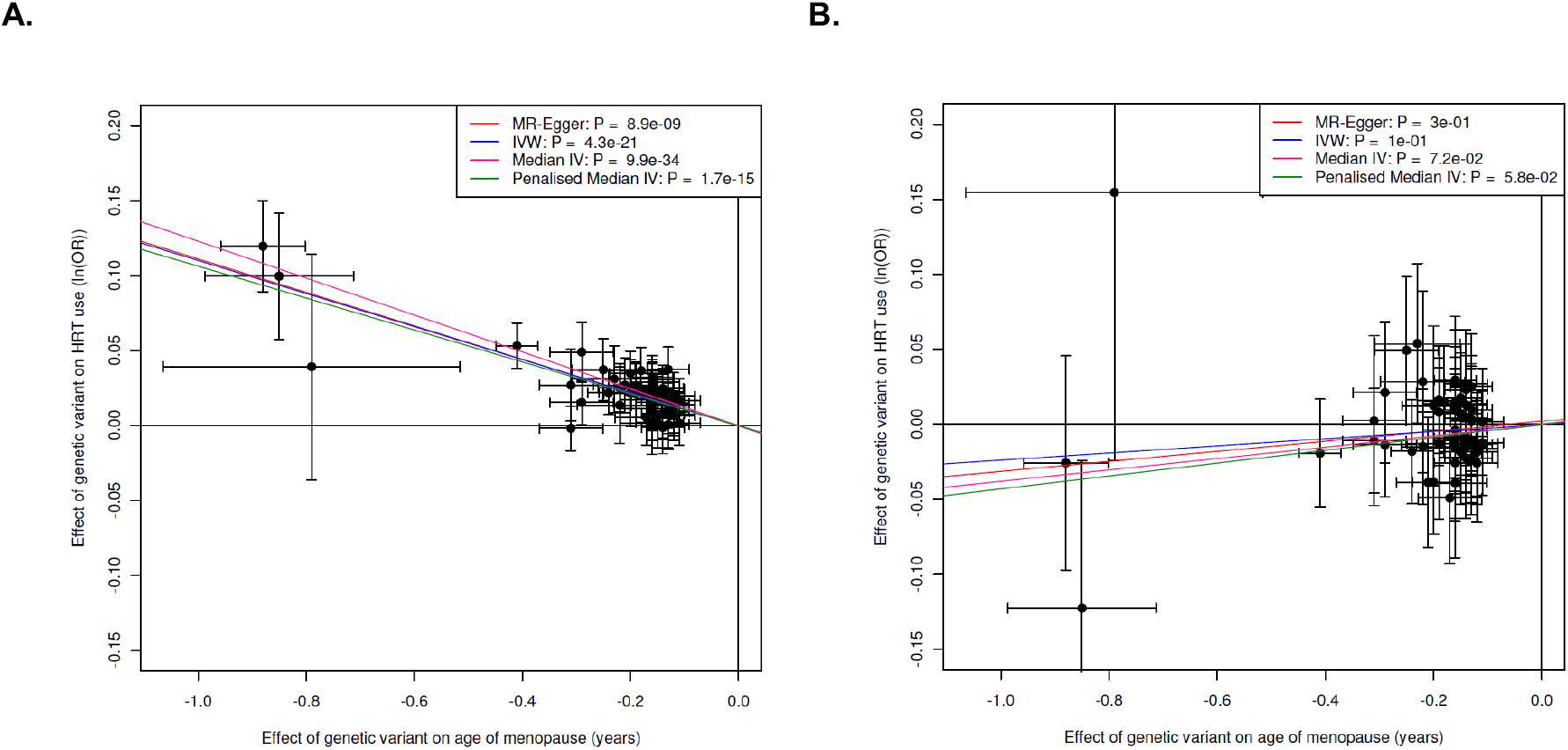
Effect of age at natural menopause on HRT use: (A) before 2002; and (B) after 2002. Mendelian randomisation analyses showing effect of genetic instrument for age at menopause on odds of using HRT.

## Discussion

In this large study of menopausal VMS, we provide further evidence to support the role of *TACR3* in the genetic basis of VMS.^15^ We identified considerable phenotypic and genetic heterogeneity at the *TACR3* locus that should be explored further to provide important insight into potential therapies targeted at *TACR3*. Our analyses provide support for using HRT as a proxy phenotype for VMS, suggesting the possibility for expanded future GWAS analyses as this phenotype may be more available than that of VMS in genetic studies. However, we show that genetic associations with therapeutic phenotypes need to be considered in the context of any major changes in the use of that therapy in routine clinical practice. Based on our results we suggest that HRT use post-2002 is more appropriate for capturing severe menopausal symptoms than pre-2002 use.

We replicated a genetic signal in *TACR3* associated with VMS^15^, adding to genetic evidence for a role of NK3R (coded for by *TACR3*) in VMS. In postmenopausal women, oestrogen withdrawal results in hypertrophy of kisspeptin-neurokinin B-dynorphin secreting (KNDy) neurons in the infundibular nucleus of the hypothalamus, which increase mRNA expression of NKB, dynorphin, kisspeptin, substance P and ERα.^42^ Studies in rodents have shown that increased levels of NKB result in more signalling through NK3R, stimulating pre-optic thermoregulatory areas of the hypothalamus but do not entirely exclude contributions of other KNDy peptides to thermoregulation (i.e. dynorphin, kisspeptin and substance P).^3,4^ In humans, previous studies have demonstrated that infusion of NKB in pre-menopausal women induces menopausal VMS^40^ and, in clinical trials, that NK3R antagonists reduce VMS in menopausal women^5,39,40^.providing support for *TACR3* as the most likely causal gene in the GWAS locus. However, our *in silico* analyses found no additional evidence to confirm *TACR3* as the causal gene and we identified no reduction in VMS in 41 heterozygous carriers of a rare loss-of-function genetic variant in *TACR3* (rs144292455)^34,43^ in whom NK3R expression should be reduced. Thus we cannot rule out the possibility that *TACR3* is not the causal gene for the VMS GWAS signal and that the GWAS signal is tagging a variant with an effect on another gene. Other published GWAS studies have identified significant signals in *TACR3* associated with testosterone levels (in males, and males and females combined)^44^, and signals near *TAC3* (which codes for NKB) as associated with age at menopause^41^. Further expanded genomic analyses of VMS and related phenotypes and the increasing availability of additional genomic data will allow these inconsistencies to be explored.

We provide evidence that the mechanism through which the genetic variants in *TACR3* influence VMS are distinct from effects of variants in this genomic region on puberty timing, consistent with known biology supporting distinct pathways. At puberty, NKB signalling is involved in the activation of Kiss1 neurons, resulting in pulsatile secretion of kisspeptin and consequently gonadotropin releasing hormone and luteinising hormone^10^. The rare variant in *TACR3* tested in our analyses (rs144292455) causes a premature stop codon (p.W275X) in the fifth transmembrane segment of the 465 amino acid NK3R and is predicted to be pathogenic and cause loss of function.^34,43^ Variant rs144292455 causes idiopathic hypogonadotropic hypogonadism in male homozygotes^34^ and the rare allele shows additive effects, delaying menarche by 1·25 years in female heterozygotes^13,35^, an effect confirmed in our study cohort despite the lack of association with VMS. One explanation for these apparently contradictory results could be that complete inhibition of NK3R signalling^45^ might be required to reduce VMS, and that the heterozygous women in our analyses still had sufficient NKB/NK3R signalling to cause VMS, resulting in no discernible reduction in symptoms. In contrast, we suggest that puberty timing appears to be sensitive to the amount of NKB/NK3R signalling. We were unable to test the effects of homozygous loss-of-function of rs144292455 on VMS, as there were no such women in our study cohort. Common genetic variation associated with age at menarche at the *TACR3* locus is independent of rare variant rs144292455^13^ and was also not associated with VMS. We propose that changes in gene regulation and expression mediate the VMS phenotype, potentially through alterations in transcription factor binding, rather than the direct effects on NK3R, and that this should be explored in future studies.

We demonstrate an interaction between a major change in clinical practice and genetic determinants for HRT use, adding to the relatively small number of robust gene-by-environment interactions in the literature. Overall, women with earlier menopause were more likely to use HRT. Prior to 2002, earlier age at menopause increased the odds of HRT use but this relationship did not exist after 2002. In contrast, the genetic signal for VMS use in *TACR3* (rs34867104) remained associated with using HRT across the time period and was not associated with menopause timing. We suggest that prior to 2002, as recommended at the time, women took HRT as a routine treatment to relieve menopausal symptoms and also to prevent complications secondary to oestrogen depletion, for example, reduced bone mineral density. After 2002, we postulate that only women with severe VMS took HRT due to perceived risks that led to an aversion of women, health professionals and clinical guidelines to HRT.^46^ It is surprising that earlier age of menopause was not associated with menopausal timing after 2002, as typically HRT will still be recommended until the average natural age of menopause for women with an early age of menopause. The lack of association of age at menopause with HRT use post 2002 highlights a potential under usage of HRT in the UK. Current UK National Institute for Health and Care Excellence guidance is that HRT should be considered as a treatment for VMS^47^ and our analysis suggests that HRT may now be underutilised among such women. Our findings suggest that genetic associations with HRT use that do not change over time are likely to be within the causal pathway for VMS.

Our analyses were carried out in Europeans in UK Biobank, in which there are known biases towards healthy and more affluent individuals^48^, highlighting a need for replication in other cohorts and ethnicities. The impact of changes to clinical practice regarding HRT use may not be the same in all populations and future analyses must be mindful of other such gene by environment interactions. Currently, linked primary care data in UK Biobank are limited to approximately half the cohort and the coding of VMS depends on women consulting a GP with symptoms, and on these being recorded. Inclusion of women with VMS without a GP record as controls in our analyses will have resulted in reduced statistical power, but not false positives. The proxy phenotype of taking HRT did not distinguish between types of medication and is based on retrospectively collected questionnaire data, which may be subject to recall bias. However, another published study that analysed self-reported medication data in UK Biobank identified the same genetic signal for HRT use.^49^

By identifying genetic associations with VMS and with the proxy phenotype HRT use we replicated a genetic signal for VMS in *TACR3* and identified a further two signals that require replication. We identified significant gene-by-time interactions for HRT use associated with menopause timing but not VMS, providing insights into the factors driving treatments and allowing refinement of a proxy phenotype for VMS. Despite the strong evidence for a role of TACR3 in VMS from clinical trials of NK3R antagonist treatments, our study highlights limitations in current understanding that should be addressed in future studies to further benefit the development of therapies for these common, potentially life-changing symptoms.

## Supporting information

Supplementary Methods, Results and Figures

## Data Availability

Data used in the present study are available on application to UK Biobank.

## Data availability

Genome-wide summary statistics will be made available at https://www.ebi.ac.uk/gwas/

## Author contributions

AM and KSR designed the study. KSR led the data analysis with contributions from RNB, JML, JT, GH and ARW. AM, KSR, RNB, JT, GH, ARW, MNW and TMF had access to the underlying UK Biobank data. CJC provided data for the VMS meta-analysis. KSR, JKP, KAP, TMF, MNW and AM reviewed and interpreted the results. KSR and AM wrote the first draft of the report. All authors reviewed the drafts and contributed to the revision of the report.

## Acknowledgements

This research was conducted using the UK Biobank resource under application number 871. KSR is supported by Cancer Research UK [grant number C18281/A29019]. JKP is supported by the UKRI Expanding Excellence in England award.

