## Supplementary Methods, Results and Figures for "Genomic insights into the mechanism of NK3R antagonists for treatment of menopausal vasomotor symptoms"

**Contents:**

p2 Supplementary Methods

p5 Supplementary Results

p6 References

p7 Supplementary Figures

### Supplementary Methods

#### Phenotype definitions

*Vasomotor symptoms derived from primary care records*

We identified Read v2 and CTV3 codes containing the full and partial words “flush”, “flash”, “meno”, “sweat”, “vaso” in the term descriptions, to account for the two clinical coding systems used in the UK Biobank linked primary care data. This resulted in the identification of 970 code and term descriptions which were manually reviewed along with their frequency of use in females in UK Biobank, resulting in the identification of 11 Read v2 and 22 CTV3 codes that were considered to be of direct relevance for vasomotor menopausal symptoms (VMS) (Supplementary Table 1). We used primary care data linked to UK Biobank to identify women with one or more clinical events containing codes for VMS. Such clinical events are coded from interactions with health care professionals in general practice including consultations, diagnoses, history, symptoms, procedures, laboratory tests and secondary care interactions reported back to general practice.

*Proxy phenotype definitions*

For secondary analyses we derived proxy phenotypes for VMS based on self-reported hormone replacement therapy use. These proxy phenotypes included the binary phenotype “Ever taken HRT” in postmenopausal women (n=72,535 cases and n=80,617 controls), and the quantitative traits “Age started HRT”, “Age ended HRT” and “Time taken HRT” in up to 85,488 women (Supplementary Table 2). We derived phenotypes based on the most recent study visit with non-missing information as described below. In the phenotype calculations, ages were rounded to ages in years.

<u>HRT ever</u>

We included women who answered “Yes” to “2724: Have you had your menopause (periods stopped)?”. Cases (used HRT) answered “Yes” to “2814: Have you ever used hormone-replacement therapy (HRT)?” while controls answered “No”. Women were excluded if they provided conflicting information at separate UK Biobank visits.

<u>HRT before and after 2002</u>

The year in which the woman started HRT was calculated from year of birth plus the age at which HRT was started. Women using HRT before 2002 were those who were ‘HRT ever’ cases and who started taking HRT before or during 2002, while those using HRT after 2002 were ‘HRT ever’ cases who started taking HRT after 2002. Controls were as described for ‘HRT ever’.

<u>Age started HRT</u>

We included women who gave an age for “3536: How old were you when you first used HRT?” that was younger than their age at the respective study visit and who answered “Yes” to “2814: Have you ever used hormone-replacement therapy (HRT)?”.

Age ended HRT

We included women who gave an age for “3546: How old were you when you last used HRT?” that was younger than their age at the respective study visit and who answered “Yes” to “2814: Ever used hormone-replacement therapy (HRT)?”.

Length of time that HRT was taken

We calculated the difference between age starting and ending HRT in women with this information available who answered “Yes” to “2814: Have you ever used hormone-replacement therapy (HRT)?”, provided that the age ending HRT was after that starting HRT.

**Calculation of proportion of women using HRT**

To calculate the proportion of women taking HRT, we identified women who stated that they were postmenopausal at baseline and thus ‘at risk’ of taking HRT in 2006−2010. To compare the proportion of women using HRT before and after 2002, we used year of birth to identify which of these postmenopausal women turned 50 (the average age of menopause in the UK) before and after 2002 (i.e. born before 1952 or in/after 1952) to minimise the effect of differences in age distribution in the cohort on temporal trends in HRT use.

**Genome-wide association study analyses**

*Age used for adjustment of statistical analyses*

Statistical analyses were adjusted for the age of the woman which was calculated at the study visit used to define the phenotype.

*Fisher’s exact test of the associations for HRT use*

Since mixed linear models can result in inflation of test statistics at low allele frequencies and adjustment for covariates can contribute to this, for genome-wide significant signals identified for case−control phenotypes we carried out a Fisher’s exact test. We calculated the total number of effect and other alleles in the case and control groups for the genetic signals and tested the null hypothesis of no difference in the distribution using a Fisher’s exact chi squared test carried out in Stata-MP v14. We considered genome-wide significant associations to be confirmed if $P<5\times10^{-8}$.

*Matched case−control sensitivity analyses*

To ensure that different age distributions of the cases and controls did not affect the effect estimates for HRT use, we generated effect estimates from case−control matched analyses. For the phenotypes ever used HRT, used HRT before 2002 and used HRT after 2002, cases and controls were 1:1 matched on age and year of birth using “ccmatch” in Stata-MP v16. This produced 51,628 age matched case/control pairs for ever used HRT, 44,431 for used HRT before 2002 and 5,615 for used HRT after 2002. Effect estimates were generated for the genetic variants of interest using case-control matched logistic regression (“clogit”) in Stata including the covariates genotyping chip and release of genotype data, recruitment centre, age and the first five genetic principal components.

**Exome-wide analyses**

We carried out gene burden association testing of rare variants using exome sequencing data available in ~200,000 people from UK Biobank [1]. Detailed sequencing methodology is provided by Szustakowski *et al.*[1] Briefly, exomes were captured with the IDT xGen Exome Research Panel v1.0 which targeted 39Mbp of the human genome with coverage exceeding on average 20x on 95.6% of sites. The OQFE protocol was used for mapping and variant calling to the GRCh38 reference. Variants included in our analyses had individual and variant missingness <10%, Hardy Weinberg Equilibrium p-value $>10^{-15}$, minimum read depth of 7 for SNPs and 10 for indels, and at least one sample per site passed the allele balance threshold > 15% for SNPs and 20% for indels.

### Supplementary Results

#### *In silico* functional and expression analyses

Two variants in strong LD with rs34867104 ($r^2$>0.8) were expression quantitative trait loci (eQTLs) for *TACR3* in brain in PsychENCODE; alleles raising *TACR3* expression were associated with lower odds of HRT use (FDR=$3\times10^{-2}$; Supplementary Table 9), an apparently opposite effect to that expected from the clinical studies of NK3R antagonism. However, these variants were not the top eQTL for *TACR3* in the region and the strongest eQTL for TACR3 in brain (rs3846440, chr4:104840863, FDR=$8\times10^{-24}$) was only weakly correlated with the lead GWAS signal for VMS (LD $r^2$<0.01 with rs34867104), and is likely to represent a distinct causal variant for gene expression. The VMS lowering allele of the lead variant for VMS (rs34867104) was associated with lower levels of methylation at a probe overlapping the promoter region of *TACR3* (P=$2\times10^{-118}$) in goDMC suggesting higher expression, though this was not the top variant associated with methylation of this probe (top variant P<$1\times10^{-300}$). The contradictory expression, methylation and clinical study evidence suggests a complex biological mechanism resulting in VMS.

#### GWAS associations with VMS and age at menarche at *TACR3*

Known genetic signals for age at menarche in/near the *TACR3* gene were not associated with VMS (*P*>0.02 for all) and showed little correlation with the VMS signal, rs34867104 (LD $r^2$≤0.12 for all) (Figure 1). The VMS signal rs34867104 was associated with age at menarche at P<$5\times10^{-8}$ in the most recent published GWAS (0.07 years per AT allele, *P*=$1.2\times10^{-10}$) (Figure 1).[2] However, in our UK Biobank analyses, conditioning on the genotypes of the menarche signals attenuated the association of rs34867104 with age at menarche (*P*=$7.5\times10^{-16}$ in single variant analyses; *P*=$2.9\times10^{-6}$ in conditional analyses) but not VMS (*P*=$3.6\times10^{-16}$ in single variant analyses; *P*=$2.3\times10^{-16}$ in conditional analyses) (Supplementary Table 5), suggesting that the association of rs34867104 with age at menarche is being driven by residual linkage disequilibrium. Taken together with the LOF analyses, our results suggest that age at menarche and VMS have a different underlying genetic architecture at the *TACR3* locus.

#### Genome-wide analyses of HRT phenotypes

We carried out genome-wide analyses of HRT use in UK Biobank as a proxy phenotype for VMS. Our GWAS included up to 153,152 women for the binary phenotype “Ever taken HRT”, derived to include postmenopausal women only (Supplementary Methods and Supplementary Table 2). In addition, we analysed the quantitative traits “Age started HRT”, “Age ended HRT” and “Time taken HRT”. In total we observed 21 signal−trait pairs at P<$5\times10^{-8}$ across the four phenotypes that following confirmation by Fisher’s exact test (binary traits; Supplementary Methods), resolved to 15 independent genetic signals across phenotypes on the basis of distance and linkage disequilibrium (Supplementary Table 6, Supplementary Figure 2).

### Supplementary Figures

**Supplementary Figure 1. Quantile-quantile and Manhattan plots for the GWAS of VMS in UK Biobank.** The plots include all variants with imputation quality>0.3 and MAF>0.001

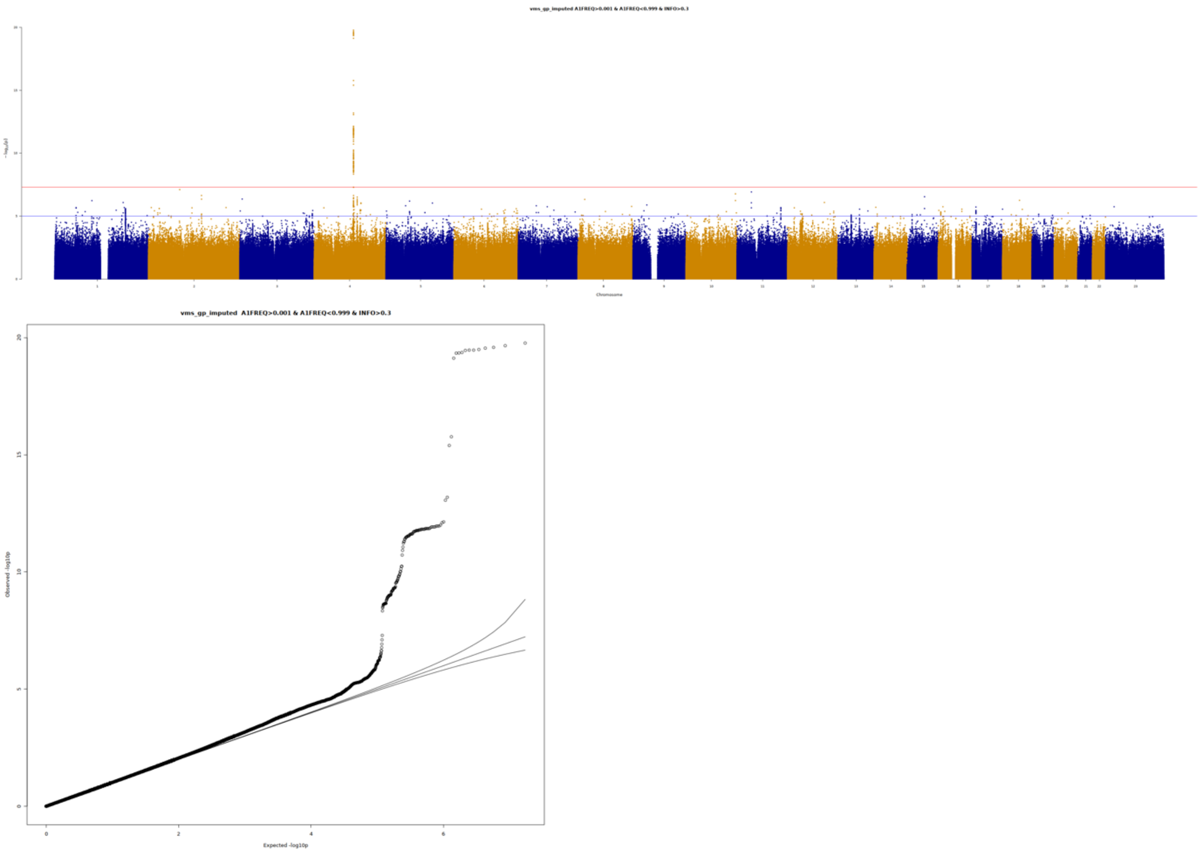

**Supplementary Figure 2. Quantile-quantile and Manhattan plots of the GWASs of HRT use.** The plots include all variants with imputation quality>0.3 and MAF>0.001. (a) HRT ever. Of the five signals identified as P<5E-08 in our GWAS, only the top SNP (rs34867104) on chromosome 4 had Fisher's exact P<5E-08; (b) HRT start age; (c) HRT end age; (d) Time taken HRT

*(a)*

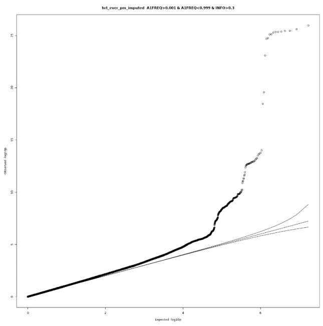

*Lambda-GC=1.09.*

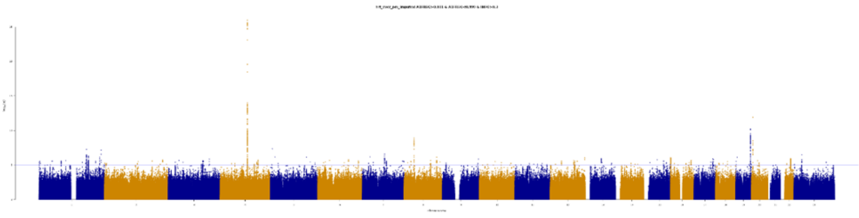

*(b)*

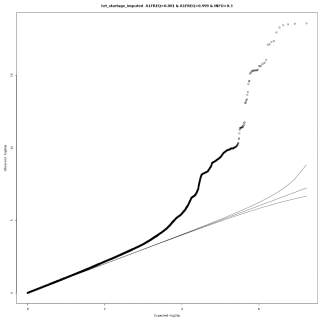

*Lambda-GC=1.05*

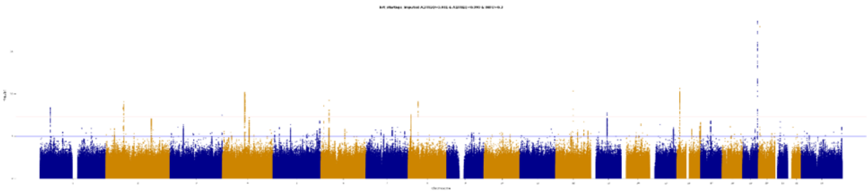

*(c)*

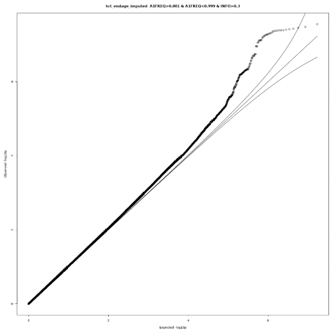

*Lambda-GC=1.05*

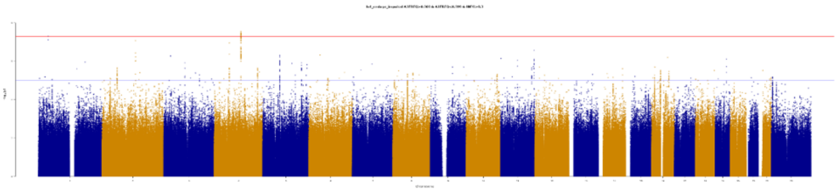

*(d)*

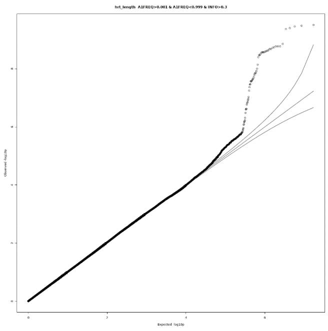

*Lambda-GC=1.00*

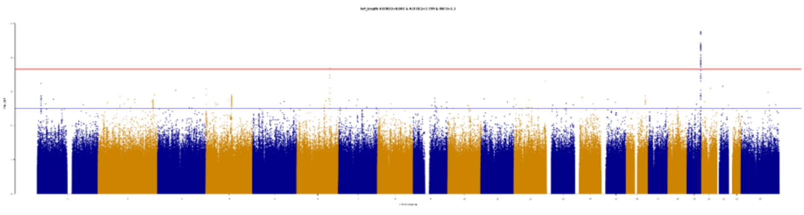
